# Mental health during the COVID-19 pandemic in two longitudinal UK population cohorts

**DOI:** 10.1101/2020.06.16.20133116

**Authors:** Alex S. F. Kwong, Rebecca M. Pearson, Mark J. Adams, Kate Northstone, Kate Tilling, Daniel Smith, Chloe Fawns-Ritchie, Helen Bould, Naomi Warne, Stan Zammit, David Gunnell, Paul Moran, Nadia Micali, Abraham Reichenberg, Matthew Hickman, Dheeraj Rai, Simon Haworth, Archie Campbell, Drew Altschul, Robin Flaig, Andrew M. McIntosh, Deborah A. Lawlor, David Porteous, Nicholas J. Timpson

**Author notes:** Corresponding Author: Alex S. F. Kwong, Population Health Sciences, Bristol Medical School, University of Bristol, United Kingdom., E. Contributed equally to this work (Alex S. F. Kwong, Rebecca M. Pearson). Contributed equally to this work (David Porteous, Nicholas J. Timpson).

## Abstract

**Background:** The impact of COVID-19 on mental health is unclear. Evidence from longitudinal studies with pre pandemic data are needed to address (1) how mental health has changed from pre-pandemic levels to during the COVID-19 pandemic and (2), whether there are groups at greater risk of poorer mental health during the pandemic?

**Methods:** We used data from COVID-19 surveys (completed through April/May 2020), nested within two large longitudinal population cohorts with harmonised measures of mental health: two generations of the Avon Longitudinal Study of Parents and Children (ALPSAC): the index generation ALSPAC-G1 (n= 2850, mean age 28) and the parent’s generation ALSPAC-G0 (n= 3720, mean age = 59) and Generation Scotland: Scottish Family Health Study (GS, (n= 4233, mean age = 59), both with validated pre-pandemic measures of mental health and baseline factors. To answer question 1, we used ALSPAC-G1, which has identical mental health measures before and during the pandemic. Question 2 was addressed using both studies, using pre-pandemic and COVID-19 specific factors to explore associations with depression and anxiety in COVID-19.

**Findings:** In ALSPAC-G1 there was evidence that anxiety and lower wellbeing, but not depression, had increased in COVID-19 from pre-pandemic assessments. The percentage of individuals with probable anxiety disorder was almost double during COVID-19: 24% (95% CI 23%, 26%) compared to pre-pandemic levels (13%, 95% CI 12%, 14%), with clinically relevant effect sizes. In both ALSPAC and GS, depression and anxiety were greater in younger populations, women, those with pre-existing mental and physical health conditions, those living alone and in socio-economic adversity. We did not detect evidence for elevated risk in key workers or health care workers.

**Interpretation:** These results suggest increases in anxiety and lower wellbeing that may be related to the COVID-19 pandemic and/or its management, particularly in young people. This research highlights that specific groups may be disproportionally at risk of elevated levels of depression and anxiety during COVID-19 and supports recent calls for increasing funds for mental health services.

**Funding:** The UK Medical Research Council (MRC), the Wellcome Trust and University of Bristol.

## Introduction

The coronavirus disease 2019 (COVID-19) pandemic has resulted in radical changes to societies globally. As the number of infected cases and deaths increased, many countries adopted public health measures including lockdown, social distancing and self-isolation. While such measures may be important for reducing transmission, they may also have a profound effect on mental health, [1-3] However, the extent to which mental health is affected by COVID-19 and its management, and who is at greatest risk, is unknown. Longitudinal studies with pre-pandemic data are vital for addressing this.

Although not directly comparable to COVID-19, evidence from previous viral outbreaks provide relevant information. The Severe Acute Respiratory Syndrome epidemic (SARS) resulted in public health mitigation measures in some countries and was associated with an increase (from pre-pandemic levels) in anxiety, depression, suicide and post-traumatic stress disorder during and beyond the conclusion of the outbreak for survivors of the virus, [4-6] and the unexposed public. [7, 8] Compared to SARS, the COVID-19 pandemic is greater in scale, resulting in more infections and deaths as well as more extreme mitigation methods. The consequences for mental health resulting from the COVID-19 pandemic could therefore be substantial. [1, 9] Unlike previous outbreaks, COVID-19 is widespread meaning the impact on global economy and health could be unprecedented.

Several rapid cross-sectional surveys during the COVID-19 pandemic have suggested a higher prevalence of anxiety, depression, [10, 11] and low wellbeing compared to historical estimates. [12, 13] However, these studies lack pre-pandemic information in the same people, reporting symptoms during and before the pandemic. This precludes accurate assessment of changes in mental health. Furthermore, selection (due to mental health influencing who respond to surveys) and reporting bias (those who perceive depression and anxiety as higher or are more likely to report symptoms when they feel there is a ‘valid’ reason) could threaten the validity of results from cross-sectional surveys. [14] There is a need for data from longitudinal designs with well-characterized sampling frames and pre-pandemic data. Such studies can more accurately identify changing patterns of mental health and identify risk groups, informing development of interventions for those at heightened risk and aiding policy decisions regarding the immediate management of COVID-19, including plans for easing restrictions, as well as for the longer-term care for groups whose mental health may be particularly affected. [1, 9]

The COVID-19 pandemic is likely to exacerbate existing social and psychological inequalities. [15] Previous studies have identified several groups who may be at greater risk of poorer mental health during COVID-19, including younger people, women, healthcare workers and those with poorer financial or living circumstances. [10, 11, 13, 16] Parents with school-aged children, individuals at risk of physical and emotional abuse and those at greater physical risk of COVID-19 (older age, and those with chronic conditions such as, asthma, obesity, diabetes) may also be at heightened risk of poorer mental health.

We used data from two large longitudinal cohort studies, both with rich pre-pandemic measures of mental health, to address (1) how has mental health changed from pre-pandemic levels to the COVID-19 pandemic and (2) are there groups at greater risk of poorer mental health during the pandemic? The first of these is important for exploring the impact of COVID-19 and its management on mental health and potential increases in poor mental health long-term. The second is important for targeting of mental health care needs now and during any subsequent waves and for identifying groups who might benefit from long-term monitoring after the pandemic.

## Methods

### Samples

We selected two comparable cohort studies to allow replication in different regions of the UK, but with similar timings of mental health measures before and during the COVID-19 pandemic.

The Avon Longitudinal Study of Parents and Children (ALSPAC) is an ongoing longitudinal population-based study that recruited pregnant women residing in Avon in the south-west of England with expected delivery dates between 1^st^ April 1991 and 31^st^ December 1992. [17, 18] The cohort consists of 13,761 mothers and their partners (hereafter referred to as ALSPAC-G0), and their 14,901 children (ALSPAC-G1). [19] The study website contains details of all data available through a fully searchable data dictionary (http://www.bristol.ac.uk/alspac/researchers/our-data/). Ethical approval for the study was obtained from the ALSPAC Ethics and Law Committee and the Local Research Ethics Committees.

Generation Scotland: Scottish Family Health Study (GS) is a family longitudinal study of 24,084 individuals recruited across Scotland between 2006 and 2011.[20] Participants were recruited into the study if they were aged 18 or over. Participants of GS have been followed up longitudinally, [21] and further details can be found on the study website (http://www.generationscotland.org). Ethical approval for the study was approved by NHS Tayside Committee on Medical Research Ethics (reference 05/S1401/89).

This study uses data from 3720 ALSPAC-G0 and 2973 ALSPAC-G1 who completed an online questionnaire about the impact and consequences of the COVID-19 pandemic between 9^th^ April and 14^th^ May 2020 (see, appendix figure 1 and figure 2) [22]. In GS, data were from 4,233 individuals who completed a similar online COVID-19 questionnaire between 17^th^ April and 17^th^ May 2020 (see, appendix figure 3). Lockdown was announced in the UK on the 24^th^ March.

### COVID-19 pandemic measures of mental health

The measures used in the COVID-19 survey examine symptoms in the preceding 2 weeks, thus represent mental health in the immediate period following lockdown. Depressive symptoms in ALSPAC were measured using the Short Mood and Feelings Questionnaire (SMFQ), [23] a 13-item instrument examining depressive mood. Scores range between 0-26 with higher scores indicting higher depressive symptoms. In GS, depressive symptoms were measured using the Patient Health Questionnaire (PHQ-9), [24] a 9-item instrument which monitors depressive symptoms. Scores range between 0-27 with higher scores indicating worse depressive symptoms. Anxiety symptoms in ALSPAC and GS was measured using the same instrument, the Generalised Anxiety Disorder Assessment (GAD-7), [25] a 7-item instrument which measures the presence of generalised anxiety disorder symptoms. Scores range between 0-21 with higher scores indicting higher anxiety symptoms. Mental wellbeing in ALSPAC and GS was also measured using the same instrument, the Short Warwick-Edinburgh Mental Wellbeing Scale (SWEMWBS), [26] a 7-item instrument which measures positive mental wellbeing. Scores range between 7-35, with higher scores indicating better mental wellbeing. These measures have recommended cut-offs for examining the proportion of individuals with probable depression (≥11 on SMFQ and ≥10 on PHQ-9), generalised anxiety disorder (≥10 on GAD-7) and poor mental wellbeing (≤17 on SWEMWBS), with good sensitivity and specificity for identifying clinical disorder using validated interviews and instruments and widely used in primary care and clinical trials (see appendix methods). Herein we refer to depressive symptoms as depression and anxiety symptoms as anxiety.

### Baseline (pre-pandemic) measures of mental health and factors

Baseline depression and anxiety were assessed in ALSPAC and GS prior to the COVID-19 pandemic. In ALSPAC-G1, baseline mental wellbeing was also assessed. These measures are described in Table 1, alongside information on the baseline factors that may predict poorer mental health in COVID-19. We refer to these as factors to make it clear that we are not assuming they are causal but could be useful for identifying vulnerable groups. The factors we explored included demographic and social information such as sex, age, educational background, financial circumstances, deprivation status, victimisation and being a parent with school-aged children. Additional factors included pre-existing mental health conditions, substance misuse, genetic risk for depression, cognitive styles, personality traits and difficulties accessing mental health information. Due to differences in data collection, several factors are only assessed in either ALSPAC or GS. We also examined associations with several COVID-19 specific factors that may be valuable predictors of adverse mental health, including obesity, asthma, infection status, isolation status, living alone, access to a garden, health-care worker and key worker status. Detailed information regarding the descriptions and timing of these measures including availability and how measures were harmonised, are given in the appendix method section.

**Table 1.** Baseline mental health measures and factors assessed in ALSPAC and Generation Scotland.

|  | ALSPAC-G0 | ALSPAC-G1 | Generation Scotland |
| --- | --- | --- | --- |
|  | Measure | Measure | Measure |
| **<u>Sociodemographic factors</u>** |  |  |  |
| Sex | Questionnaire item | Questionnaire item | Questionnaire item |
| Age | Questionnaire item | Questionnaire item | Questionnaire item |
| Educational background | Questionnaire item | Questionnaire item | Questionnaire item |
| Income | Questionnaire item | Questionnaire item | Questionnaire item |
| Deprivation status | Indexes of multiple deprivation | Indexes of multiple deprivation | Indexes of multiple deprivation |
| Recent financial problems | Questionnaire item | Questionnaire item | Questionnaire item |
| Partner emotional abuse | Life events questionnaire | Not assessed | Not assessed |
| Parent with young children | Not assessed | Questionnaire item | Questionnaire item |
| **<u>Biological factors</u>** |  |  |  |
| Obesity | BMI > 30 assessed at a research clinic | BMI > 30 assessed at a research clinic | BMI > 30 assessed at a research clinic |
| Asthma | Questionnaire item | Questionnaire item | Questionnaire item |
| **<u>COVID-19 specific factors</u>** |  |  |  |
| Infection status | Questionnaire item | Questionnaire item | Questionnaire item |
| Isolation status | Questionnaire item | Questionnaire item | Not assessed |
| Living alone | Questionnaire item | Questionnaire item | Questionnaire item |
| No access to a garden | Questionnaire item | Questionnaire item | Questionnaire item |
| Health care worker | Questionnaire item | Questionnaire item | Not assessed |
| Key worker | Questionnaire item | Questionnaire item | Questionnaire item |
| **<u>Baseline mental health measures</u>** |  |  |  |
| Depressive symptoms | Edinburgh postnatal depression scale | Short mood and feelings questionnaire | General health questionnaire - depression |
| Anxiety | Spielberger state-trait anxiety inventory | Generalised anxiety disorder assessment | General health questionnaire - anxiety |
| Mental wellbeing | Not assessed | Warwick-Edinburgh mental wellbeing scale | Not assessed |

|  |  |  |  |
| --- | --- | --- | --- |
| Probable major depression (MDD) | Life events questionnaire | Clinical interview schedule - revised | Structured clinical interview for DSM-IV |
| Probable generalised anxiety disorder (GAD) | Life events questionnaire | Clinical interview schedule - revised | Not assessed |
| Psychosis like experiences | Not assessed | Psychosis like symptoms interview | Schizotypal personality questionnaire |
| Disordered eating | Life events questionnaire | Questionnaire items consistent with DSM-V frequency | Not assessed |
| Obsessive compulsive disorder traits (OCD) | Not assessed | Obsessive-compulsive inventory | Not assessed |
| Autistic traits | Not assessed | Social responsiveness scale | Not assessed |
| Personality disorder traits | Karolinska scales of personality | Standardised assessment of personality: abbreviated scale | Not assessed |
| History of alcohol misuse | Alcohol use disorders identification test | Alcohol use disorders identification test | Questionnaire item |
| Current smokers (tobacco) | Questionnaire items | Questionnaire items | Questionnaire items |
| Cognitive styles | Negative schemas questionnaire | Cognitive styles questionnaire | Brief resilience scale |
| Difficulties accessing mental health information | Not assessed | Public health questionnaire | Not assessed |
| Neuroticism | Not assessed | Big five factors of personality – neuroticism | Eysenck personality questionnaire – neuroticism |
| Self-harm history | Not assessed | Questionnaire items | Linkage to medical records |
| Depression polygenic risk score (PRS) | Constructed from a recent GWAS on depression | Constructed from a recent GWAS on depression | Constructed from a recent GWAS on depression |

### Statistical analysis

Analysis was conducted in StataSE (version 15). Initially, we described the prevalence of mental health outcomes during the COVID-19 pandemic in all cohorts. To answer our first research question, we used ALSPAC-G1 to examine how mental health changed from baseline (pre-pandemic) to COVID-19 levels. This analysis was only possible in ALSPAC-G1 who had identical mental health measures at baseline and during COVID-19.

To answer our second research question, we examined associations between factors (both baseline measures and those specific to and measured during the pandemic) and COVID-19 depression and anxiety. Analysis was conducted separately for all cohorts, adjusting for sex, age and the date they completed the COVID-19 questionnaire (to account for heterogeneity in response times). In ALSPAC-G0 and GS, we used alternative measures capturing the same construct to account for pre-existing depression and anxiety (the EPDS in ALSPAC-G and the GHQ-28 in GS). Wellbeing was not assessed at baseline in ALSPAC-G1 or GS, therefore we restrict this analysis to depression and anxiety only. The results for question two can be interpreted as identifying factors associated with depression and anxiety, that are not driven by past symptoms of either disorder (as they are adjusted for in the model). Continuous COVID-19 and baseline depression and anxiety were standardised to create Z scores allowing comparison of effect sizes across outcomes and cohorts.

#### Missing data

Our eligible samples were defined as those who completed at least one mental health measure during the COVID-19 surveys: ALSPAC-G0 n= 3579, ALSPAC-G1 n= 2872 and GS n =4208 (appendix figures 1-3, appendix table 1). We imputed incomplete baseline depression, anxiety and factors using earlier or concurrent information up to the eligible samples, using multiple imputation by chained equations to generate 50 imputed datasets. [27] Full information on the proportion of missing baseline data in each cohort are given in the appendix table 2-4, the majority (>80%) of participants had more than 50% of complete baseline data (i.e., all factors and baseline mental health) with less than 1% only having information on only 1 or 2 baseline variables. Analyses on multiple imputed datasets uses rich pre-pandemic data available to plausibly meet the assumption that data are missing at random, i.e., conditional on observed information. Whereas, complete case analyses assume that missingness is not related to the outcome conditional on the exposure and any covariables. Given that we have demonstrated that the missing baseline data was associated with lower education and sex (appendix table 1) it is likely that the complete case sample is biased and thus we primarily present imputed estimates which also increase power. Details regarding imputation are fully described in the appendix.

**Table 2.** Associations between baseline risk factors and depression and anxiety using the imputed samples. Results are standardised estimates for depression and anxiety, adjusted for prior depression or anxiety, sex, age and when the COVID-19 questionnaire was completed. *Indicates the % of individuals with caseness for ALSPAC-G0 / ALSPAC-G1 / GS respectively. **Indicates a continuous scale was used so no proportions are given. ***Indicates this was on complete case analysis only. MDD: major depressive disorder; GAD: generalised anxiety disorder; OCD: obsessive compulsive disorder; PRS: polygenic risk score.

|  | Depression standardised estimates (95% CIs), *P* |  |  | Anxiety standardised estimates (95% CIs), *P* |  |  |
| --- | --- | --- | --- | --- | --- | --- |
|  | ALSPAC G0<br>(n=3579) | ALSPAC G1<br>(n=2872) | Gen Scot<br>(n=4208) | ALSPAC G0<br>(n=3579) | ALSPAC G1<br>(n=2872) | Gen Scot<br>(n=4208) |
| **<u>Sociodemographic factors</u>** |  |  |  |  |  |  |
| Sex (female)<br>(76% / 72% / 64%)* | 0.28 (0.21, 0.34)<br>*P* = 1.46 x 10<sup>-19</sup> | 0.22 (0.15, 0.29)<br>*P* = 1.20 x 10<sup>-9</sup> | 0.27 (0.22, 0.32)<br>*P* = 4.07 x 10<sup>-22</sup> | 0.26 (0.20, 0.32)<br>*P* = 7.87 x 10<sup>-17</sup> | 0.40 (0.33, 0.48)<br>*P* = 3.57 x 10<sup>-23</sup> | 0.18 (0.12, 0.23)<br>*P* = 6.63 x 10<sup>-10</sup> |
| Age (older ages)<br>(scale variable)** | -0.02 (-0.03, -0.01)<br>*P* = 2.08 x 10<sup>-8</sup> | -0.01 (-0.07, 0.04)<br>*P* = 0.643 | -0.02 (-0.02, -0.02)<br>*P* = 4.57 x 10<sup>-46</sup> | -0.02 (-0.03, -0.02)<br>*P* = 4.82 x 10<sup>-12</sup> | 0.00 (-0.06, 0.07)<br>*P* = 0.945 | -0.01 (-0.02, -0.01)<br>*P* = 1.25 x 10<sup>-30</sup> |
| Lower educational<br>background<br>(11% / 16% / 13%) | 0.23 (0.12, 0.35)<br>*P* = 0.00008 | 0.05 (-0.04, 0.15)<br>*P* = 0.276 | 0.15 (0.06, 0.24)<br>*P* = 0.001 | 0.14 (0.03, 0.25)<br>*P* = 0.016 | 0.16 (0.06, 0.26)<br>*P* = 0.002 | 0.09 (0.00, 0.18)<br>*P* = 0.061 |
| Higher income<br>(scale variable) | -0.04 (-0.06, -0.03)<br>*P* = 6.22 x 10<sup>-8</sup> | -0.01 (-0.04, 0.01)<br>*P* = 0.328 | -0.10 (-0.13, -0.08)<br>*P* = 5.14 x 10<sup>-16</sup> | -0.01 (-0.03, 0.00)<br>*P* = 0.052 | -0.04 (-0.07, -0.01)<br>*P* = 0.008 | -0.07 (-0.09, -0.04)<br>*P* = 5.46 x 10<sup>-7</sup> |
| Worse deprivation status<br>(scale variable) | 0.07 (0.04, 0.10)<br>*P* = 0.00002 | 0.01 (-0.01, 0.04)<br>*P* = 0.319 | 0.06 (0.04, 0.09)<br>*P* = 7.33 x 10<sup>-7</sup> | 0.05 (0.02, 0.08)<br>*P* = 0.001 | 0.04 (0.01, 0.07)<br>*P* = 0.005 | 0.04 (0.02, 0.06)<br>*P* = 0.001 |
| Financial problems<br>(11% / 10% / 4%) | 0.29 (0.15, 0.43)<br>*P* = 0.00005 | 0.14 (0.03, 0.26)<br>*P* = 0.015 | 0.38 (0.18, 0.58)<br>*P* = 0.0002 | 0.20 (0.07, 0.32)<br>*P* = 0.011 | 0.24 (0.12, 0.36)<br>*P* = 0.0002 | 0.20 (0.00, 0.39)<br>*P* = 0.049 |
| Partner emotional abuse<br>(8% / Na / Na) | 0.36 (0.18, 0.53)<br>*P* = 0.00005 | Not assessed | Not assessed | 0.29 (0.13, 0.46)<br>*P* = 0.001 | Not assessed | Not assessed |
| Parent with young<br>children<br>(Na / 11% / 11%) | Not assessed | 0.03 (-0.07, 0.13)<br>*P* = 0.570 | -0.03 (-0.14, 0.08)<br>*P* = 0.617 | Not assessed | 0.19 (0.08, 0.30)<br>*P* = 0.001 | 0.05 (-0.05, 0.16)<br>*P* = 0.353 |
| **<u>Biological factors</u>** |  |  |  |  |  |  |
| Obesity<br>(18 % / 14% / 20%) | 0.22 (0.12, 0.32)<br>*P* = 0.00001 | 0.18 (0.06, 0.31)<br>*P* = 0.004 | 0.32 (0.24, 0.40)<br>*P* = 1.09 x 10<sup>-14</sup> | 0.14 (0.04, 0.25)<br>*P* = 0.005 | 0.15 (0.03, 0.27)<br>*P* = 0.012 | 0.10 (0.02, 0.18)<br>*P* = 0.010 |
| Asthma<br>(16% / 10% / 10%) | 0.07 (-0.02, 0.16)<br>*P* = 0.127 | 0.08 (-0.04, 0.20)<br>*P* = 0.196 | 0.18 (0.07, 0.28)<br>*P* = 0.001 | 0.07 (-0.02, 0.16)<br>*P* = 0.14 | 0.21 (0.07, 0.35)<br>*P* = 0.003 | 0.12 (0.02, 0.22)<br>*P* = 0.016 |
| **<u>COVID-19 specific factors</u>** |  |  |  |  |  |  |
| COVID-19 infection<br>(12% / 16% / 8%) | 0.18 (0.07, 0.28)<br>*P* = 0.001 | 0.09 (0.00, 0.17)<br>*P* = 0.045 | 0.17 (0.05, 0.29)<br>*P* = 0.004 | 0.16 (0.06, 0.27)<br>*P* = 0.003 | 0.08 (-0.02, 0.17)<br>*P* = 0.112 | 0.10 (-0.02, 0.21)<br>*P* = 0.101 |
| Self-isolation<br>(19% / 25% / Na) | 0.20 (0.11, 0.29)<br>*P* = 6.64 x 10<sup>-6</sup> | 0.15 (0.08, 0.22)<br>*P* = 0.00004 | Not assessed | 0.13 (0.04, 0.27)<br>*P* = 0.003 | 0.17 (0.09, 0.25)<br>*P* = 0.00003 | Not assessed |
| Living alone<br>(8% / 6% / 16%) | 0.45 (0.30, 0.59)<br>$P = 4.80 \times 10^{-9}$ | 0.20 (0.06, 0.34)<br>$P = 0.005$ | 0.19 (0.11, 0.27)<br>$P = 4.47 \times 10^{-6}$ | -0.06 (-0.19, 0.07)<br>$P = 0.372$ | 0.06 (-0.08, 0.21)<br>$P = 0.392$ | -0.03 (-0.11, 0.06)<br>$P = 0.539$ |
| No access to a garden<br>(2% / 18% / 8%) | 0.47 (0.09, 0.85)<br>$P = 0.016$ | 0.16 (0.07, 0.24)<br>$P = 0.0002$ | 0.24 (0.12, 0.37)<br>$P = 0.0001$ | -0.07 (-0.35, 0.21)<br>$P = 0.62$ | 0.05 (-0.04, 0.14)<br>$P = 0.235$ | 0.16 (0.04, 0.28)<br>$P = 0.007$ |
| Health care worker<br>(11% / 12% / NA) | 0.01 (-0.09, 0.10)<br>$P = 0.901$ | -0.02 (-0.12, 0.08)<br>$P = 0.683$ | Not assessed | -0.03 (-0.13, 0.07)<br>$P = 0.597$ | 0.02 (-0.08, 0.13)<br>$P = 0.652$ | Not assessed |
| Key worker<br>(32% / 39% / 22%) | 0.04 (-0.03, 0.11)<br>$P = 0.214$ | -0.09 (-0.15, -0.02)<br>$P = 0.008$ | -0.05 (-0.13, 0.02)<br>$P = 0.178$ | 0.03 (-0.04, 0.10)<br>$P = 0.441$ | 0.02 (-0.05, 0.09)<br>$P = 0.631$ | 0.04 (-0.03, 0.12)<br>$P = 0.266$ |
| **<u>Psychiatric or mental health factors</u>** |  |  |  |  |  |  |
| Probable MDD<br>(8% / 14% / 14%) | 0.38 (0.22, 0.54)<br>$P = 2.29 \times 10^{-6}$ | 0.31 (0.20, 0.42)<br>$P = 3.18 \times 10^{-8}$ | 0.39 (0.29, 0.49)<br>$P = 1.84 \times 10^{-13}$ | 0.26 (0.11, 0.40)<br>$P = 0.0005$ | 0.49 (0.39, 0.62)<br>$P = 1.33 \times 10^{-16}$ | 0.27 (0.17, 0.38)<br>$P = 7.03 \times 10^{-7}$ |
| Probable GAD<br>(7% / 13% / Na) | 0.26 (0.11, 0.42)<br>$P = 0.001$ | 0.14 (0.03, 0.25)<br>$P = 0.010$ | Not assessed | 0.25 (0.09, 0.40)<br>$P = 0.002$ | 0.50 (0.39, 0.62)<br>$P = 2.72 \times 10^{-17}$ | Not assessed |
| Psychosis like experiences<br>(Na / 15% / scale) | Not assessed | 0.17 (0.07, 0.27)<br>$P = 0.001$ | 0.15 (0.11, 0.19)<br>$P = 3.72 \times 10^{-14}$ | Not assessed | 0.25 (0.15, 0.36)<br>$P = 4.74 \times 10^{-6}$ | 0.12 (0.08, 0.16)<br>$P = 9.59 \times 10^{-9}$ |
| Disordered eating<br>(3% / 9% / Na) | 0.09 (-0.14, 0.32)<br>$P = 0.689$ | 0.21 (0.09, 0.34)<br>$P = 0.0005$ | Not assessed | 0.08 (-0.16, 0.32)<br>$P = 0.510$ | 0.26 (0.12, 0.40)<br>$P = 0.0002$ | Not assessed |
| OCD traits<br>(scale variable) | Not assessed | 0.05 (0.01, 0.09)<br>$P = 0.027$ | Not assessed | Not assessed | 0.15 (0.11, 0.19)<br>$P = 8.28 \times 10^{-13}$ | Not assessed |
| Autistic traits<br>(Na / 7% / Na) | Not assessed | 0.19 (0.05, 0.34)<br>$P = 0.008$ | Not assessed | Not assessed | 0.35 (0.20, 0.51)<br>$P = 5.18 \times 10^{-6}$ | Not assessed |
| Personality disorder traits<br>(11% / 11% / Na) | 0.32 (0.19, 0.45)<br>$P = 1.67 \times 10^{-6}$ | 0.09 (0.04, 0.23)<br>$P = 0.169$ | Not assessed | 0.15 (0.02, 0.27)<br>$P = 0.021$ | 0.27 (0.14, 0.40)<br>$P = 0.00008$ | Not assessed |
| History of alcohol misuse<br>(17% / 9% / 16%) | 0.04 (-0.05, 0.13)<br>$P = 0.367$ | 0.13 (0.01, 0.25)<br>$P = 0.040$ | 0.02 (-0.06, 0.10)<br>$P = 0.598$ | 0.09 (0.00, 0.18)<br>$P = 0.047$ | 0.20 (0.07, 0.33)<br>$P = 0.003$ | -0.02 (-0.10, 0.06)<br>$P = 0.569$ |
| Current smokers (tobacco)<br>(29% / 12% / 10%) | 0.18 (0.10, 0.25)<br>$P = 7.58 \times 10^{-6}$ | 0.02 (-0.09, 0.13)<br>$P = 0.690$ | 0.30 (0.19, 0.41)<br>$P = 1.23 \times 10^{-7}$ | 0.12 (0.05, 0.20)<br>$P = 0.001$ | 0.10 (-0.01, 0.21)<br>$P = 0.085$ | 0.18 (0.08, 0.29)<br>$P = 0.001$ |
| Negative cognitive styles<br>(scale variable) | 0.21 (0.16, 0.25)<br>$P = 1.07 \times 10^{-18}$ | 0.09 (0.05, 0.13)<br>$P = 0.00004$ | 0.22 (0.19, 0.26)<br>$P = 2.97 \times 10^{-32}$ | 0.16 (0.12, 0.20)<br>$P = 3.07 \times 10^{-14}$ | 0.07 (0.02, 0.12)<br>$P = 0.003$ | 0.19 (0.15, 0.22)<br>$P = 5.39 \times 10^{-21}$ |
| Difficulties accessing mental health info (Na / 23% / Na) | Not assessed | 0.12 (0.03, 0.20)<br>*P* = 0.009 | Not assessed | Not assessed | 0.28 (0.19, 0.36)<br>*P* = 1.93 x 10<sup>-9</sup> | Not assessed |
| Higher neuroticism (scale variable) | Not assessed | 0.04 (0.01, 0.09)<br>*P* = 0.015 | 0.22 (0.19, 0.26)<br>*P* = 3.00 x 10<sup>-42</sup> | Not assessed | 0.11 (0.07, 0.15)<br>*P* = 7.33 x 10<sup>-7</sup> | 0.21 (0.18, 0.25)<br>*P* = 1.97 x 10<sup>-31</sup> |
| Self-harm history (Na / 24% / 2%) | Not assessed | 0.15 (0.06, 0.23)<br>*P* = 0.001 | 0.55 (0.22, 0.88)<br>*P* = 0.001 | Not assessed | 0.19 (0.09, 0.28)<br>*P* = 0.0002 | 0.58 (0.28, 0.88)<br>*P* = 1.97 x 10<sup>-8</sup> |
| Depression PRS*** (scale variable) | 0.09 (0.05, 0.13)<br>*P* = 0.00002<br>(n=1906) | 0.03 (-0.02, 0.07)<br>*P* = 0.224<br>(n=1592) | 0.05 (0.02, 0.08)<br>*P* = 0.0002<br>(n=3849) | 0.09 (0.05, 0.14)<br>*P* = 0.00004<br>(n=2071) | 0.00 (-0.05, 0.05)<br>*P* = 0.993<br>(n=1329) | 0.06 (0.03, 0.09)<br>*P* = 0.00002<br>(n=3832) |

#### Sensitivity analyses

In sensitivity analysis, we also examined depression and anxiety using the ‘proportion-above-threshold’ in logistic regressions rather than continuous scores as outcomes, analysed varying ages for baseline measures and estimated ‘counterfactual’ trajectories for the mental health measures to highlight differences in the observed compared to predicted trajectories.

## Results

Data on COVID-19 mental health outcomes in ALSPAC-G0 were available for 3579 people (mean age: 58.67 years, SD: 4.82), for ALSPAC-G1, 2872 people (mean age: 27.61 years, SD: 0.54) and GS 4208 people (mean age: 59.24 years, SD: 12.03), see appendix figures 1-3, appendix table 1.

### Prevalence of mental health outcomes during COVID-19

The prevalence of probable depression decreased with age in ALSPAC and GS. Similar results were observed for probable anxiety and low wellbeing (Figure 1).

**Figure 1.**
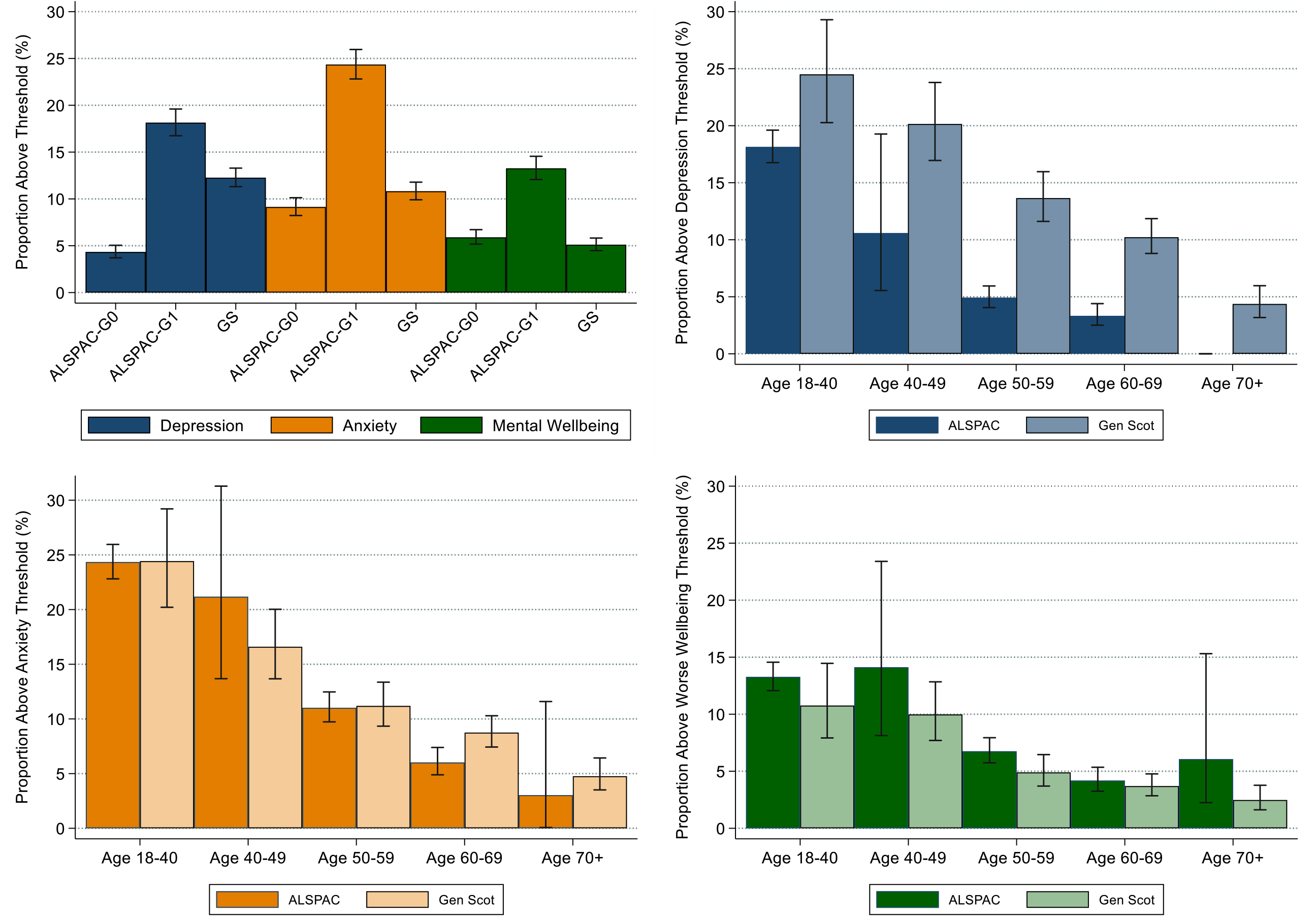
Mental health during COVID-19 in ALSPAC-G0, ALSPAC-G1 and Generation Scotland (GS). Figure 1A (top left) shows probable depression, probable generalised anxiety disorder (GAD) and lower wellbeing by each cohort. Figure 1B (top right) shows probable depression by age groups assessed using the SMFQ in ALSPAC and PHQ-9 in GS. Figure 1C (bottom left) shows probable GAD by age groups assessed by the GAD-7. Figure 1D shows lower wellbeing by age groups assessed by the SWEMWBS. Note, ALSPAC-G1 (n=2812) were categorised as 18-40, even though the max age of this cohort is 29 years. Age in ALSPAC-G0 was split by the following: Age 40-49 (n=89), Age 50-59 (n=2105), Age 60-69 (n=1455) and Age 70+ (n=71). In GS, Age was split by the following: Age 18-40 (n=356), Age 40-49 (n=534), Age 50-59 (n=964), Age 60-69 (n=1526) and Age 70+ (n=853).

### Change in mental health in ALSPAC-G1

The percentage of individuals with probable depression was lower, 18.14 % (16.76, 19.61), in COVID, compared to 24.35 % (23.04, 25.70) at the most recent baseline. The percentage of people with probable anxiety disorder was almost double during COVID-19 :24% (95% CI 23%, 26%) compared to pre-pandemic levels (13%, 95% CI 12 %, 14%) and for lower wellbeing :13, (95% CI 12 to 14%%) compared to 8% (95% CI 7 to 9%) (Figure 2, appendix table 5 and 6). When examining continuous measures of mental health, there was a mean difference in SMFQ score of −0.60 (95% CI: −0.84, −0.37), 1.36 (95% CI: 1.10, 1.61) for GAD-7 and 2.45 (95% CI 2.25 to 2.65) for SWEMWBS, when comparing the most recent baseline to COVID-19. To give a summary of magnitude, these estimates represent a 0.11, 0.26 and 0.51 standardised effect difference respectively (appendix table 7). Item level differences for each measure are shown in figure 3.

**Figure 2.**
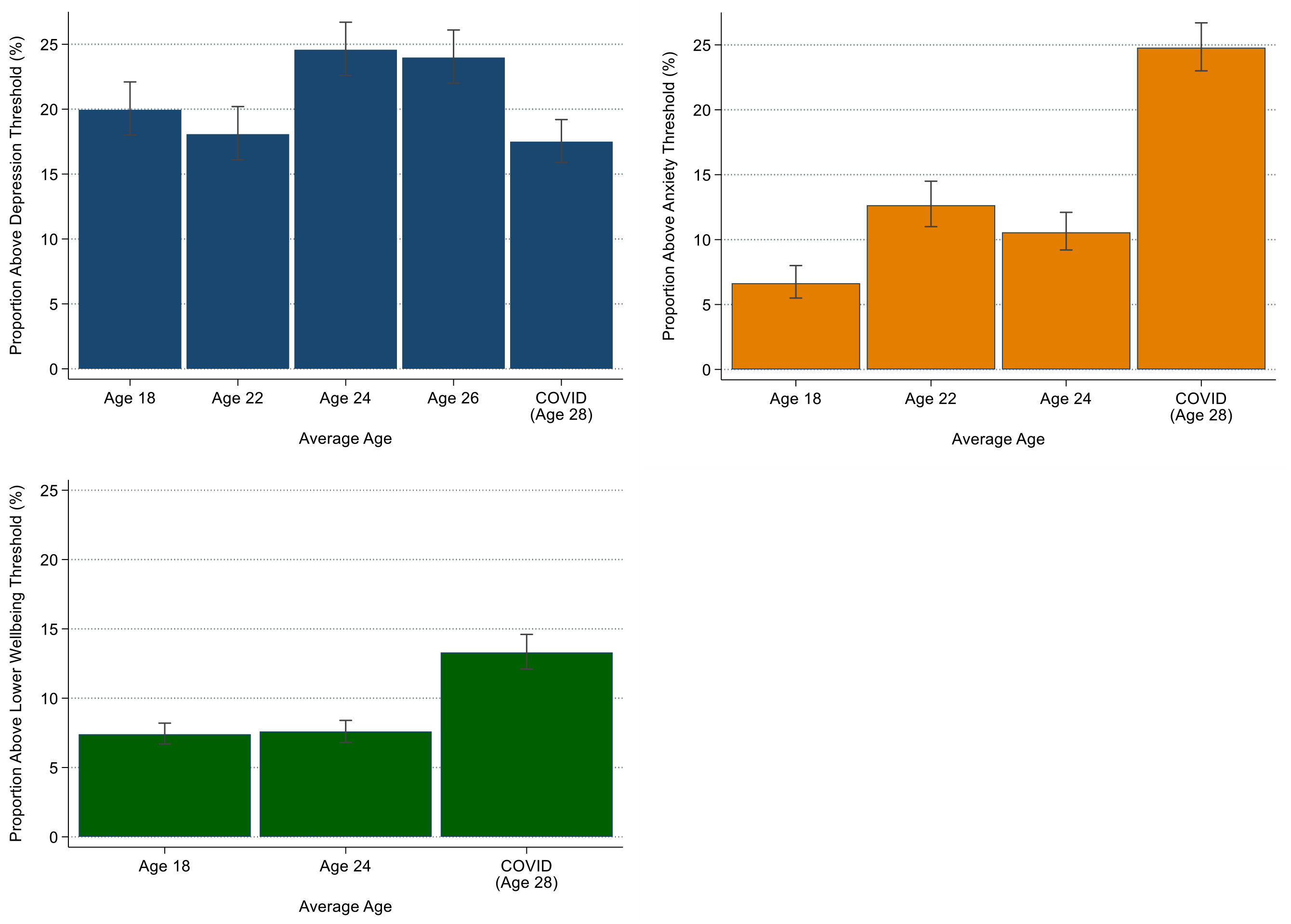
Changes in mental health across baseline (pre-pandemic) to COVID-19 in ALSPAC-G1. Figure 2A (top left) shows changes in probable depression as assessed by the SMFQ. Figure 2B (top right) shows changes in probable GAD assessed by the GAD-7 at age 22 and CISR GAD at ages 18 and 24. Figure 2C (bottom left) shows changes in lower wellbeing assessed by the SWEMWBS.

**Figure 3.**
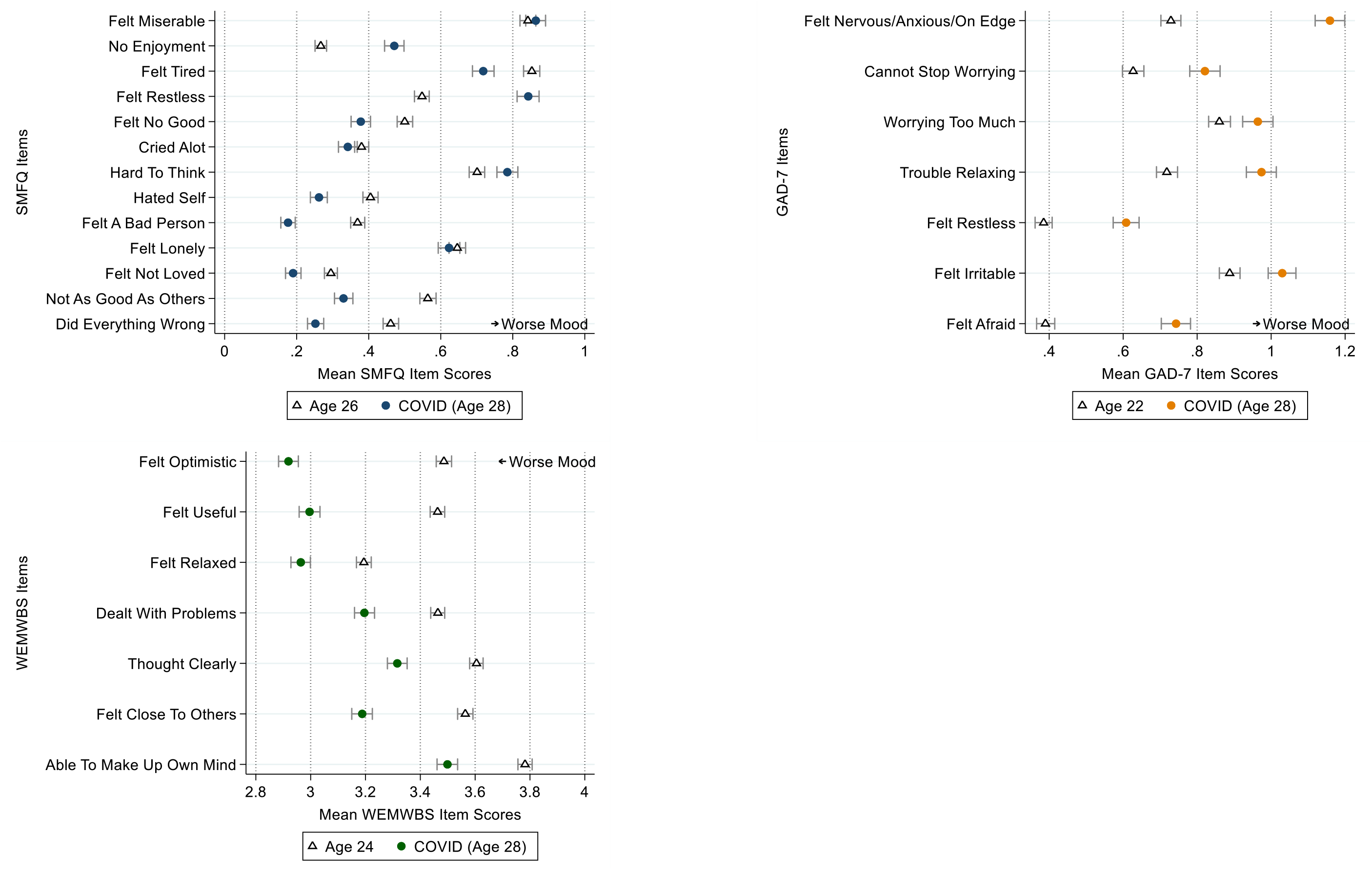
Item level changes in mental health between the most recent baseline and COVID-19 in ALSPAC-G1. Figure 3A (top left) shows how items of the SMFQ (depression) vary from the most recent baseline (Age 26) to COVID-19. Figure 3B (top right) shows how items of the GAD-7 (anxiety) vary from the most recent baseline (Age 22) to COVID-19. Figure 3C (bottom left) shows high items from the SWEMWBS (mental wellbeing) vary from the most recent baseline (Age 24) to COVID-19.

### Factors related to depression and anxiety during COVID-19

Table 2 and figures 4 and 5 show the associations between baseline and COVID-19 specific factors and depression and anxiety, with adjustment for baseline depression and anxiety symptoms (measured on a continuous scale in standard deviation units for ease of comparison between cohorts and outcomes).

**Figure 4.**
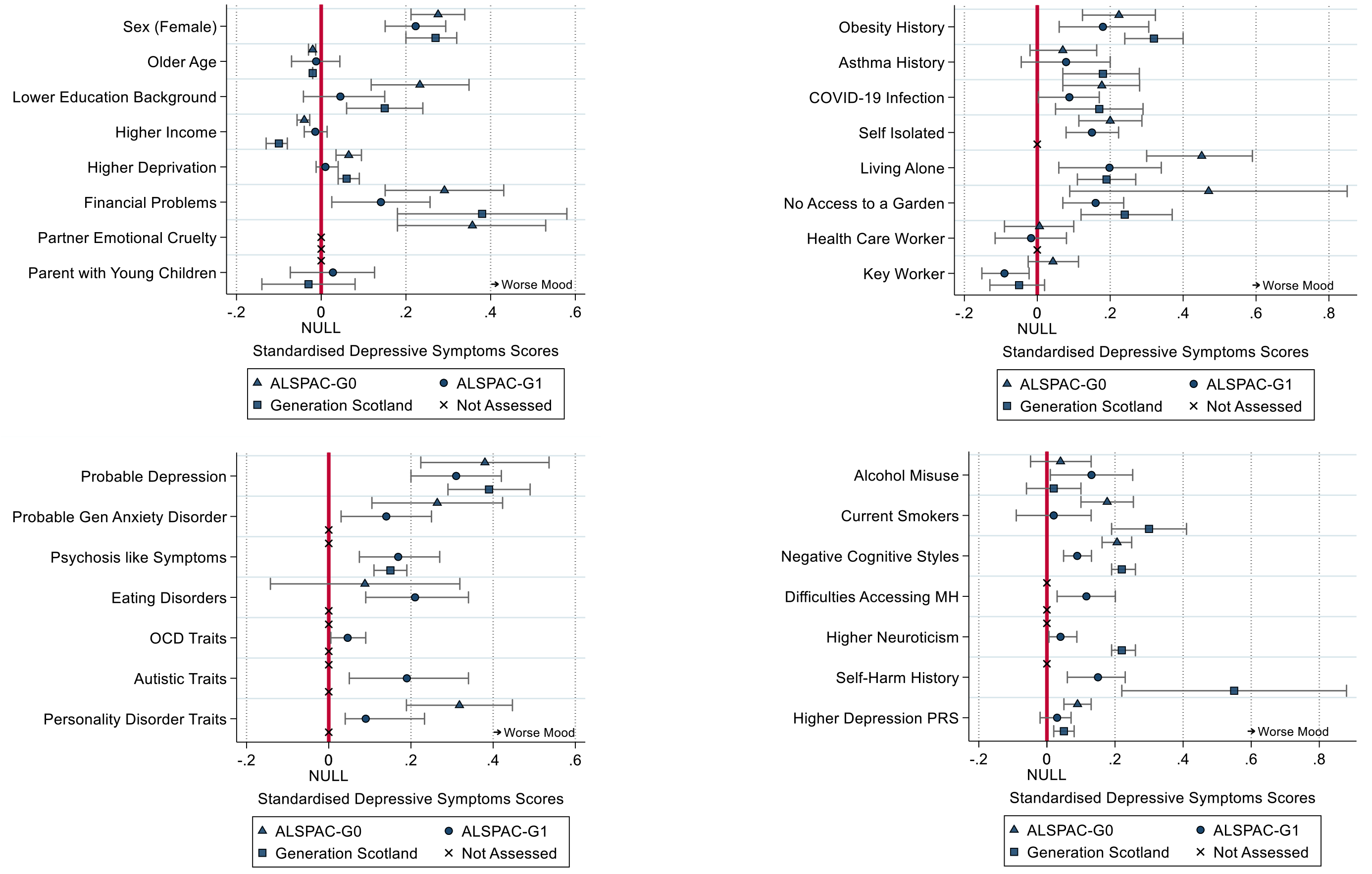
Associations between baseline and COVID-19 factors and depression during COVID-19, adjusted for baseline depression, sex, age and when the COVID-19 questionnaire was completed, using imputed data. Estimates refer to a standard deviation increase in depression, over and above depression at baseline. Figure 4A (top left) shows associations between baseline sociodemographic factors and depression during COVID-19. Figure 4B (top right) shows associations between baseline physical health and COVID-19 specific factors and depression during COVID-19. Figure 4C (bottom left) and Figure 4D (bottom right) shows associations between baseline mental health factors and depression during COVID-19.

**Figure 5.**
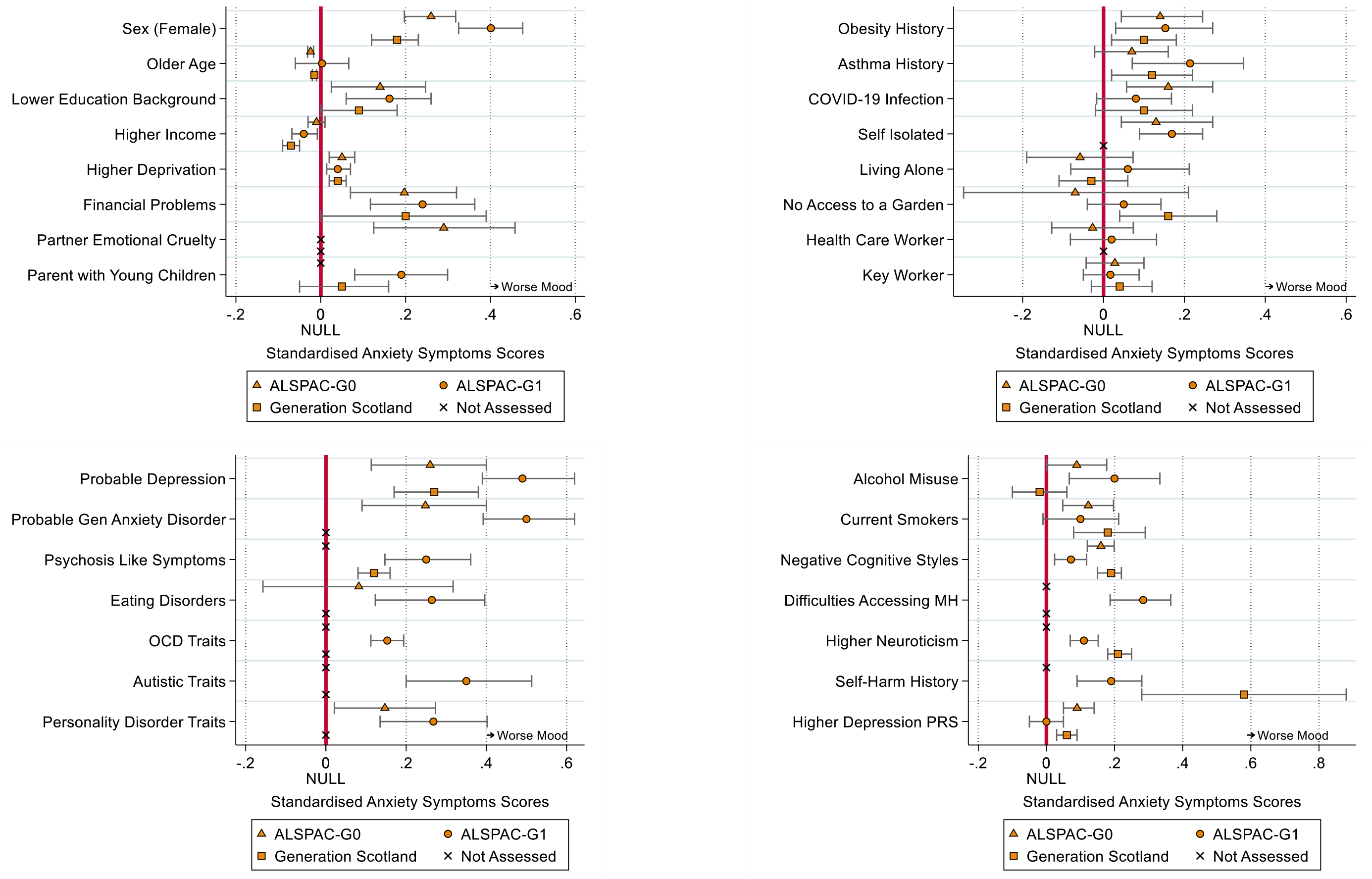
Associations between baseline and COVID-19 factors and anxiety during COVID-19, adjusted for baseline anxiety, sex, age and when the COVID-19 questionnaire was completed, using imputed data. Estimates refer to a standard deviation increase in anxiety, over and above anxiety at baseline. Figure 4A (top left) shows associations between baseline sociodemographic factors and anxiety during COVID-19. Figure 4B (top right) shows associations between baseline physical health and COVID-19 specific factors and anxiety during COVID-19. Figure 4C (bottom left) and Figure 4D (bottom right) shows associations between baseline mental health factors and anxiety during COVID-19.

A reported or suspected COVID-19 infection was associated with higher depression and anxiety in ALSPAC-G0, but only higher depression in GS with no associations observed in ALSPAC-G1. Living alone during the pandemic was associated with higher depression, but not with higher anxiety in ALSPAC and GS. No access to a garden was associated with higher depression in all cohorts and greater anxiety in GS. ALSPAC-G0 and ALSPAC-G1 participants who reported that they had self-isolated had higher depression and anxiety, but it was not possible to test this in GS. Key workers (of any kind) and health care workers were not associated with higher depression or anxiety. There was an association between being a key worker and having lower depression in ALSPAC-G1, but this was not replicated in ALSPAC-G1 or GS.

For pre-pandemic factors, focusing on replicated results (those showing consistent associations in at least two cohorts), we found evidence for higher depression and anxiety in women, those with financial problems, lower educational backgrounds, lower income, living in a more deprived area, those with obesity. A positive association of being a parent of a young child with higher anxiety in ALSPAC-G1 did not replicate in GS and was not associated with depression in either cohort.

Reporting an emotionally abusive partner was only available in ALSPAC-G0 but was positively associated with both greater depression and anxiety. Asthma had positive associations with higher anxiety in ALSPAC-G1 and GS but was not associated with depression in both cohorts.

There were strong and replicated associations between several pre-existing mental health problems and higher depression and anxiety, including a history of major depression disorder, psychosis-like symptoms, negative cognition, neuroticism, and a history of self-harm. The depression polygenic risk score was also positively associated with depression and anxiety in ALSPAC-G0 and GS (though not in ALSPAC-G1).

Depression and anxiety were higher in ALSPAC-G1 in those who reported generalised anxiety disorder, OCD traits, disordered eating, autistic traits, and difficulty accessing mental health services, but these outcomes were not available in ALSPAC-G1 or GS and so could not be replicated. Personality disorder traits were associated only with higher anxiety in G1 but both anxiety and depression ALSPAC-G0. A history of alcohol abuse was associated with increased depression in ALSAPC-G1 and anxiety in both ALSAPC-G0 and ALSPAC-G1, but not replicated in GS. By contrast smoking was associated with increased depression and anxiety in both ALSPAC-G0 and GS but not ALSPAC-G1.

Results were similar using complete case analysis (appendix table 8), adjusting for educational background with imputation (appendix table 9), using different timings of baseline depression and anxiety (appendix tables 10 and 11) and examining binary outcomes (appendix table 12).

## Discussion

We report a population-based longitudinal study to track changes in depression and anxiety from before to during the COVID-19 pandemic, providing potentially important new information for policy planning. Although we found no clear evidence that depression has changed during COVID-19 from pre-pandemic waves in ALSPAC-G1, there was strong observational evidence that anxiety and lower wellbeing were higher during COVID-19 compared to pre-pandemic levels. Approximately twice as many young adults experienced probable anxiety disorder and low wellbeing during the pandemic compared to previous waves. The mean rises of 0.26 SD in GAD-7 scores and 0.51 SD in SWEMWBS represent effect sizes that are clinically important and are seen in those following treatment (in the opposite direction). [28, 29] While mental health is dynamic and changes over time, evidence suggests that mood disorders tend to stabilise throughout adulthood, so the rise in anxiety and reduction in wellbeing in ALSPAC-G1 goes against what we would expect in the absence of COVID-19. [30] Our trajectory analyses (see appendix figure 4) suggested that higher anxiety and lower wellbeing deviated from expected levels, but that depression was in line with expectations in comparison to previous waves.

The uncertainty and sudden change to everyday life, as well as concerns over health, may explain why anxiety, rather than depression, has initially risen. The apparent rise in younger ages may reflect the impact of mitigation measures (i.e., lockdown and social distancing) rather than a risk of COVID-19 infection (which may be higher in older populations). Furthermore, depression usually relates to feelings of loss, whilst anxiety relates to threat, as the majority of participants may not yet have lost anything (e.g., death of loved one, loss of job) this may also explain why depression has currently remained stable. There is also evidence that anxiety changes more rapidly than depression following treatment. [28]. What separates this pandemic from historical outbreaks, is the global impact. This, alongside the community spirit, may have been protective against the self-blame and guilt intrinsic to depression. [31] Indeed, depression items that were lower in the pandemic, as compared to previous waves, related to feelings of self-blame. As social inequalities become apparent and threat of loss becomes actual loss, this may change. The current survey was in UK spring, whereas previous ALSPAC-G1 waves were predominantly completed in winter. Seasonal trends suggest that depression and anxiety scores are approximately 1-2 points (0.1 SD) lower and 5% less of individuals are above thresholds in spring than winter, [32] which may explain lower depression scores.

Irrespective of the overall change in depression and anxiety in each cohort, several sociodemographic, psychological, physical and COVID-19 factors were associated with greater depression and anxiety during COVID-19. A reported or suspected COVID-19 infection was a factor for higher depression and anxiety in ALSPAC-G0 and GS, possibly reflecting the high perceived risk to physical health in older ages. This supports previous research, [2] but must be interpreted with caution because COVID-19 status here largely includes participants’ perception that they have COVID-19. Therefore, it maybe that those with pre-existing depression and anxiety are more likely to perceive that their symptoms are COVID-19 rather than other conditions. There was consistent evidence in participants from ALSPAC and GS that health risk groups linked to COVID-19, such as those with obesity and to some extent asthma, had higher depression and anxiety during COVID-19, potentially reflecting concerns regarding perceived risk of infection or the impact of more stringent social distancing. There was no evidence that key workers or health workers were at greater risk of depression or anxiety, suggesting these groups are not yet experiencing difficulties. This may reflect the heterogenous group of occupations included in this group, but whilst we observed no initial change in these groups, as the situation continues, frontline health care workers may become at risk of PTSD.

Those reporting self-isolation were at higher risk of both anxiety and depression but living alone was consistently associated with greater depression only. The manifestation of depression rather than anxiety for those living alone may relate to loneliness which is amplified with physical contact restricted to within households, again reflecting depression being related to absence and loss rather than threat. Whereas, self-isolation (which in this context is related to having symptoms) may be linked to anxiety through associated threat of the virus. Parents of young children were more anxious in ALSPAC, which may reflect stress related to the sudden change in childcare provision. Financial problems, lower income and deprivation were associated with greater risk of depression and anxiety in ALSPAC-G0 and GS. Financial problems, but not lower income, was also associated with higher depression and anxiety in ALSPAC-G1. Whilst these cohorts may have different populations, replication of financial concerns highlights the potential importance of global policies to mitigate the sudden social-economic impact and ensure emergency financial measures are accessible. [16]

As expected, individuals with a history of poorer mental health across multiple domains were at greater risk of higher depressive and anxiety during COVID-19, supporting concerns raised at the beginning of this pandemic. [1, 33, 34] Personality traits such as neuroticism and negative thinking patterns are strong factors for higher depression and anxiety during COVID-19 and are modifiable with interventions which could benefit those at risk now or in future outbreaks, even if delivered remotely. [35] ALSPAC-G0 and GS participants with genetic risk for depression were associated with poorer mental health, yet these effects were much weaker in ALSPAC-G1.

There are several limitations to this work. Firstly, as the pandemic is a universal exposure, it is difficult to attribute factors to the impact of the COVID-19 pandemic specifically, with many factors likely to show an association with later depression and anxiety at any time. [31] However, using longitudinal data and methods, we were able to demonstrate that anxiety and lower wellbeing were worse during COVID-19 than expected, given the comparison between baseline and pandemic assessments, so it is unlikely these effects are not related to COVID-19. Secondly, there were heterogeneous measurements of mental health in COVID-19 specific surveys and baseline, as well as differences in the length of follow up across cohorts. This poses a challenge in inferring strong conclusions on change and specificity of findings to generations or cohorts. However, several sensitivity analyses in both cohorts and exploring different baselines reached similar conclusions. Thirdly, we were only able to assess change over the pandemic in ALSPAC-G1. Therefore, our inferences may only be relevant to young adult populations. However, given the replication between younger ages and higher rates of depression and anxiety, it is likely these effects will be observed in other studies. Fourthly, although we were able to use existing data such as educational background to predict baseline missingness and use such variables in imputation models, we did not impute further than the sample with complete COVID-19 survey data, given that data was unique. Thus, there may be issues with generalisability as respondents were more likely to be female and from higher educational backgrounds than previous ALSPAC and GS surveys. Furthermore, the meaning and interpretation of depression, anxiety and wellbeing may vary during pandemics, for example, some level of tension and fear may be adaptive and appropriate. However, our item level analysis revealed that all anxiety and mental wellbeing items were worse during COVID-19, implying a global decrease in these aspects of mood, not just for specific components. Finally, we compared multiple factors and therefore some statistical associations may have occurred as a result of chance. However, for most factors, there was evidence for replication of findings across multiple cohorts, suggesting that ‘chance’ findings are less likely.

Future work is needed to understand the mechanisms and complex interplay between baseline and COVID-19 specific factors and mental health during the COVID-19 pandemic. Future research should also consider how changes in anxiety might influence public behaviour through contact patterns and compliance with policies. Depression and anxiety, along with associated impairment should continue to be carefully monitored to forecast the long-term impact of this crisis. This can help ensure that future policies consider optimal preservation of both physical and mental health.

## Contributors

ASFK, RMP, MJA, KN, KT, AMM, DAL, DP and NJT contributed to the conception and design of the study. ASFK, RMP, MJA, KN, AC, SH, CFR, DA, RF, DS, DP and NJT contributed to the organisation of the conduct of the study. ASFK carried out the study (including acquisition of data). ASFK and MJA analysed the data. ASFK and RMP drafted the initial output. All authors contributed to the interpretation of data. All authors have read and approved the final version of the manuscript. ASFK will serve as guarantor for the contents of the paper.

## Data Availability

ALSPAC data is available to researchers through an online proposal system. Information regarding access can be found on the ALSPAC website (http://www.bristol.ac.uk/media-library/sites/alspac/documents/researchers/data-access/ALSPAC_Access_Policy.pdf).
GS:SFHS data is available to researchers on application to the Generation Scotland Access Committee. The managed access process ensures that approval is granted only to research which comes under the terms of participant consent.

## Data availability

ALSPAC data is available to researchers through an online proposal system. Information regarding access can be found on the ALSPAC website (http://www.bristol.ac.uk/media-library/sites/alspac/documents/researchers/data-access/ALSPAC_Access_Policy.pdf).

GS:SFHS data is available to researchers on application to the Generation Scotland Access Committee. The managed access process ensures that approval is granted only to research which comes under the terms of participant consent.

## Acknowledgments

The UK Medical Research Council and Wellcome (Grant Ref: 217065/Z/19/Z) and the University of Bristol provide core support for ALSPAC. A comprehensive list of grants funding is available on the ALSPAC website (http://www.bristol.ac.uk/alspac/external/documents/grant-acknowledgements.pdf). We are extremely grateful to all the families who took part in this study, the midwives for their help in recruiting them, and the whole ALSPAC team, which includes interviewers, computer and laboratory technicians, clerical workers, research scientists, volunteers, managers, receptionists and nurses. Part of this data was collected using REDCap, see the REDCap website for details https://projectredcap.org/resources/citations/). ASFK, RMP, KT, DS, SH, DAL, NJT work in or are affiliated to the MRC Integrative Epidemiology Unit which is funded by the University of Bristol and UK Medical Research Council (MC_UU_00011/3 and MC_UU_00011/6). European Research Council under the European Union’s Seventh Framework Programme, and grants 758813, MHINT and 669545 from the European Research Council Grant Agreements. This work was supported by Wellcome through the Wellcome Longitudinal Population Studies COVID-19 Secretariat and Steering Group (UK LPS COVID co-ordination, Grant Ref: 221574/Z/20/Z). This work was also supported by the NIHR Biomedical Research Centre at University Hospitals Bristol NHS Foundation Trust and the University of Bristol. Generation Scotland received core support from the Chief Scientist Office of the Scottish Government Health Directorates [CZD/16/6] and the Scottish Funding Council [HR03006], and is currently supported by the Wellcome Trust [216767/Z/19/Z]. Genotyping of the GS:SFHS samples was carried out by the Genetics Core Laboratory at the Wellcome Trust Clinical Research Facility, University of Edinburgh, Scotland, funded by the MRC and Wellcome Trust [104036/Z/14/Z]. This work has made use of the resources provided by the Edinburgh Compute and Data Facility (ECDF) (http://www.ecdf.ed.ac.uk/). NJT is a Wellcome Trust Investigator (202802/Z/16/Z), is the PI of the Avon Longitudinal Study of Parents and Children (MRC & WT 217065/Z/19/Z), is supported by the University of Bristol NIHR Biomedical Research Centre (BRC-1215-2001), the MRC Integrative Epidemiology Unit (MC_UU_00011) and works within the CRUK Integrative Cancer Epidemiology Programme (C18281/A19169). DG, PM, SZ and DR are supported by the NIHR Biomedical Research Centre at University Hospitals Bristol NHS Foundation Trust. None of the named funders influenced the study design, data collection, analyses or interpretation of results. The views expressed in this paper are those of the authors and not necessarily of any of the funders, the National Health Service or the Department of Health.

